# Mendelian randomization studies: a metric for quality evaluation

**DOI:** 10.1101/2025.01.08.25320206

**Authors:** Fiorella Rosas-Chavez, Tony R. Merriman

## Abstract

**Background:** Mendelian randomization (MR) is a genetic epidemiological method used to infer causal relationships between exposures and outcomes. Its application in hyperuricemia and gout has grown exponentially owing to the ready availability of summary statistics from genome-wide association studies and the ease of applying the two-sample MR technique. However indications of poor study quality suggest the need for systematic evaluation.

**Objective:** This study evaluated the quality of two-sample MR studies on hyperuricemia and gout and developed a scoring system to help reviewers and readers assess their quality and validity.

**Methods:** A systematic review was conducted on 86 two-sample MR studies published between 2016 and 2024. Studies were assessed using a scoring system encompassing study design, statistical methods, result interpretation, and adherence to STROBE-MR guidelines. Trends in quality over time were analyzed using regression models.

**Results:** Study quality scores ranged from 0 to 19, with a mean of 9.1 and median of 11, demonstrating wide variability. High-quality studies adhered to MR assumptions, used independent datasets, and conducted replication analyses, while lower-quality studies often failed to correct the p-value when needed, test for confounders, address dataset overlap or report study power. Despite the increased publication of MR studies, overall quality not improved over time.

**Conclusion:** There is variability in two-sample MR study quality. Our proposed scoring system offers a practical framework for evaluating MR studies, aiding researchers and clinicians in identifying robust findings while promoting higher methodological standards.

## Introduction

Gout is caused by a response of the innate immune system to monosodium urate crystals deposited in the joints of people with hyperuricemia(1). Hyperuricemia and gout are strongly comorbid with renal cardiometabolic conditions (2, 3) and also associated with various other diseases including cancer and neurological conditions (4). Hence, there is much interest in understanding causal relationships between gout, hyperuricemia and other conditions or comorbidities.

Mendelian randomization (MR) is a genetic epidemiological method that aims to assess causal relationships between an exposure and an outcome (5). Instead of the exposure itself, it uses inherited genetic variants as instrumental variables (IV). The variants are randomly assigned at conception and remain unaffected by environmental influences. This randomness, as explained by the second law of independent assortment of genes by Mendel, is what gives MR its name and makes it analogous to a randomized controlled trial, where groups are assigned randomly to different exposures. In application to urate control and gout, the "intervention" would be the inheritance of a urate-increasing or gout risk allele, and the “control group” would consist of individuals who inherit the other allele. MR analysis requires the instrumental variable to influence the outcome directly through the exposure, which is a difficult requirement to test (6).

MR has been particularly useful in challenging previous causality associations. For instance, MR has provided evidence against a causal role for high-density lipoprotein in cardiovascular disease(7) and evidence against a causal role for C-reactive protein in coronary heart disease(8), while supporting causality for low-density lipoprotein(9). In gout, MR has shown that circulating urate is not causally associated with chronic kidney disease (CKD) (10, 11), aligning with findings from randomized clinical trials that found no benefit of urate-lowering treatments on CKD progression(12, 13). MR studies have demonstrated a causal role for increased BMI and insulin resistance in hyperuricemia and gout(5, 14). An umbrella review of MR and other studies concluded that, while hyperuricemia is causal for gout and nephrolithiasis, it does not play a causal role in other disease phenotypes(5, 15).

The first MR studies used simple linear or logistic regression (5). If individual level data were available, the two-stage-least-squares method was often used, which allowed an estimate of the effect size of any causal relationship. More recently, MR studies have virtually exclusively used readily available summary statistics from genome-wide association studies (GWAS) for each of the exposure and outcome in an approach termed two-sample MR. An alternative, also using GWAS summary statistics, is a likelihood-based method where summary statistics are directly modelled with a likelihood function.

The availability of user-friendly statistical packages, for example MendelianRandomization in R(16), and freely-downloadable GWAS summary statistics from, for example, the UK Biobank (17), FinnGen (18) and the Veteran Affairs Million Veterans Program (19), has commoditized two-sample MR studies, substantially increasing the volume of publications in the literature. This creates challenges in evaluation of study quality for people without a background in genetic epidemiology. Therefore, the aim of this paper was to systematically evaluate two-sample MR studies in hyperuricemia and gout, and to provide guidelines for researchers and clinicians who may not be experts in MR to critically assess these studies. While our focus is on hyperuricemia and gout, we anticipate that our guidelines will have broad applicability in other medical fields.

## Methods

### Study Selection and Eligibility Criteria

We conducted a systematic review of Mendelian randomization (MR) studies focusing on hyperuricemia and gout as either exposure or outcome variables. We carried out a PubMed search on March 27, 2024, using the terms “Mendelian randomization urate” and “Mendelian randomization gout”. Only peer-reviewed studies were included if they used two-sample MR analysis and assessed urate or gout as an exposure or outcome variable. Studies not focused primarily on urate or gout were excluded. From an initial pool of 234 studies after full-text review, 56 studies were excluded due to being preprints, errata, commentaries, MR methodology articles, or review articles. Another 47 studies were excluded for not using two-sample MR, and 45 were excluded because urate or gout were not the primary focus. Ultimately, 86 studies met the inclusion criteria. (Figure 1)

**Figure 1.**
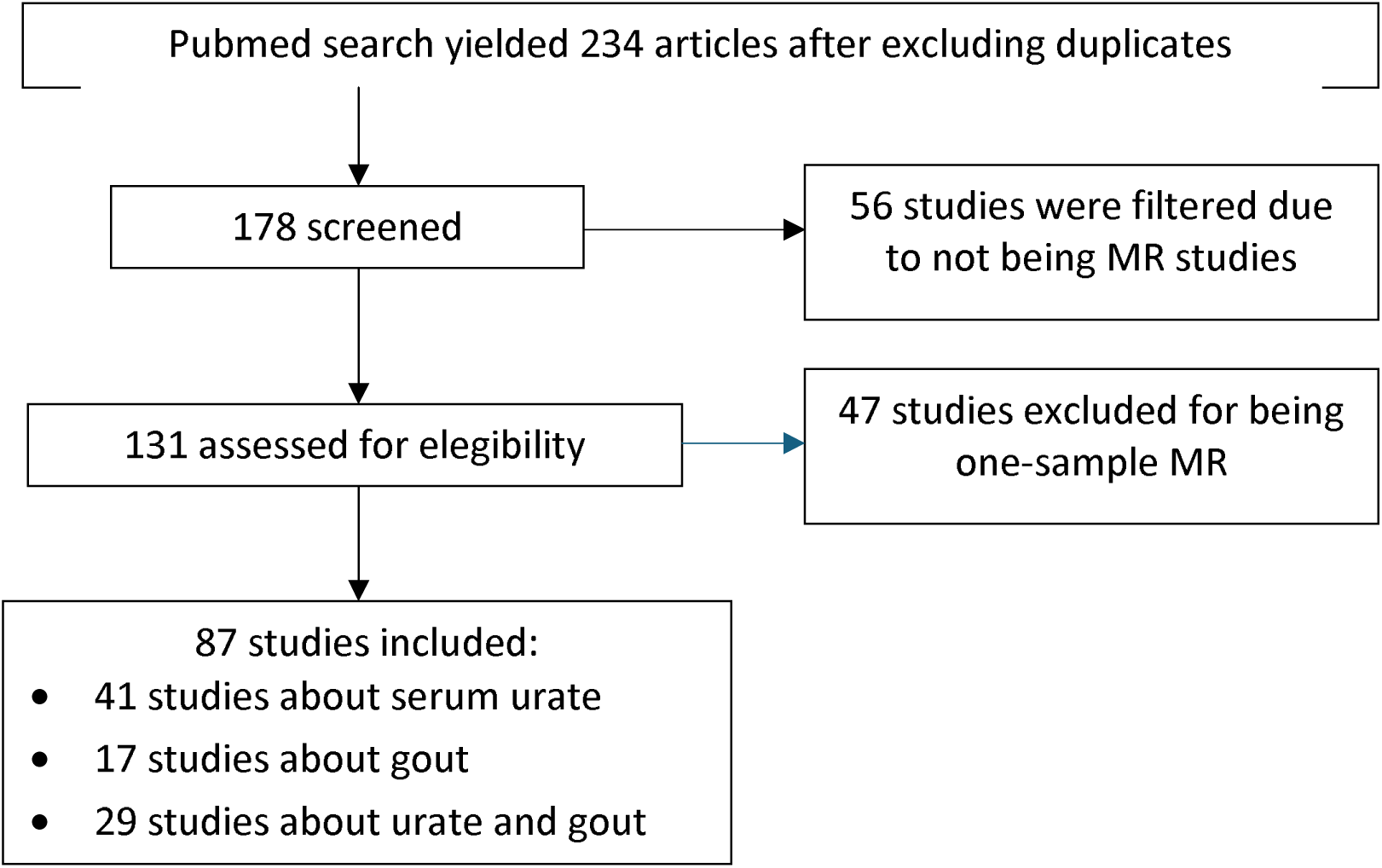
Flow diagram of review process.

### Scoring System for Study Quality Assessment

We developed a scoring system with a possible range of -9 to 21 to evaluate the studies. This system assessed factors such as study design, statistical methods, interpretation of results and adherence to STROBE guidelines (20)(Table 1). Below we describe the various factors.

**Table 1.**
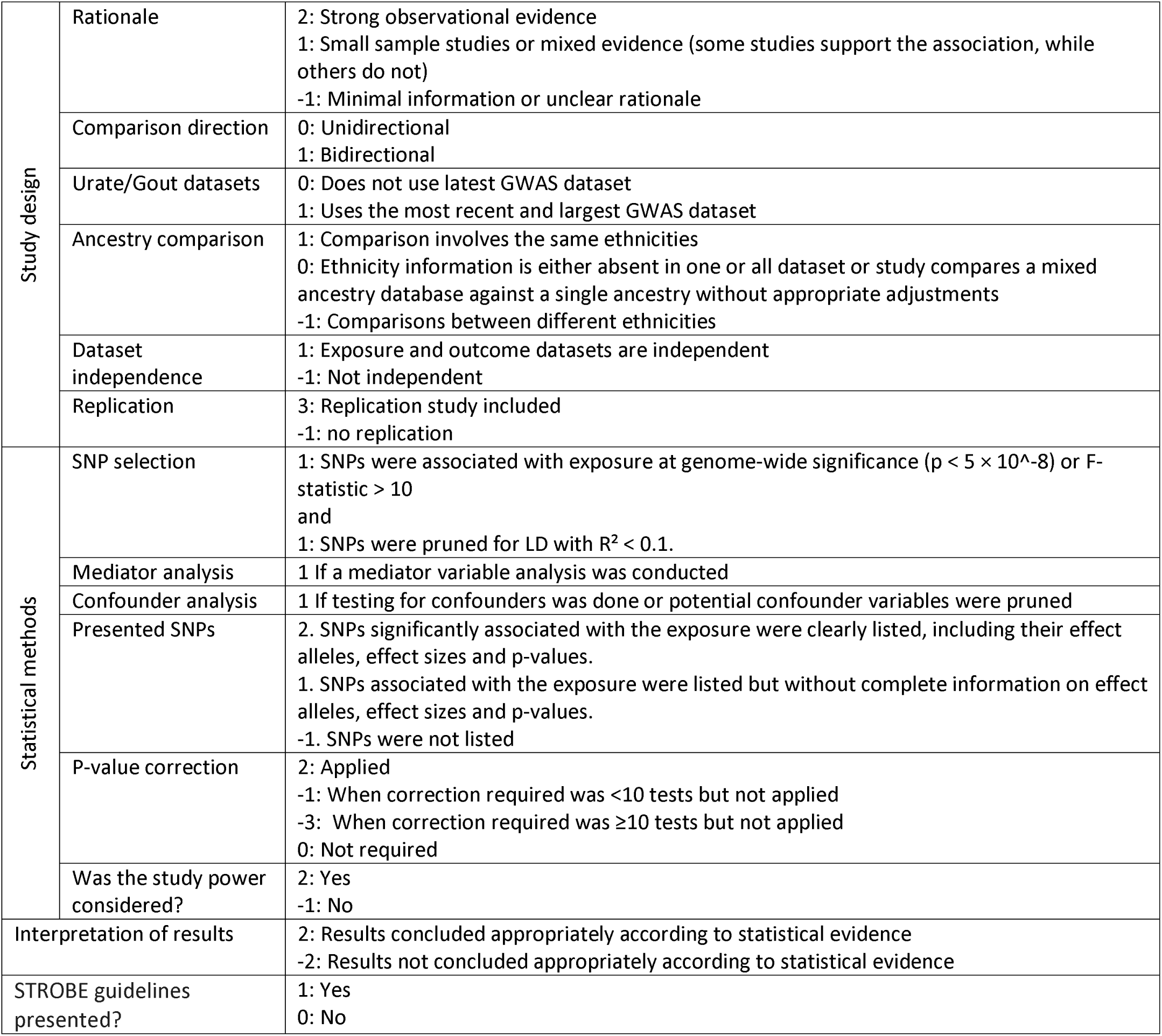
Scoring system for 2-sample MR studies.

|  |  |  |
| --- | --- | --- |
| Study design | Rationale | 2: Strong observational evidence<br>1: Small sample studies or mixed evidence (some studies support the association, while others do not)<br>-1: Minimal information or unclear rationale |
|  | Comparison direction | 0: Unidirectional<br>1: Bidirectional |
|  | Urate/Gout datasets | 0: Does not use latest GWAS dataset<br>1: Uses the most recent and largest GWAS dataset |
|  | Ancestry comparison | 1: Comparison involves the same ethnicities<br>0: Ethnicity information is either absent in one or all dataset or study compares a mixed ancestry database against a single ancestry without appropriate adjustments<br>-1: Comparisons between different ethnicities |
|  | Dataset independence | 1: Exposure and outcome datasets are independent<br>-1: Not independent |
|  | Replication | 3: Replication study included<br>-1: no replication |
| Statistical methods | SNP selection | 1: SNPs were associated with exposure at genome-wide significance ($p < 5 \times 10^{-8}$) or F-statistic $> 10$<br>and<br>1: SNPs were pruned for LD with $R^2 < 0.1$. |
|  | Mediator analysis | 1 If a mediator variable analysis was conducted |
|  | Confounder analysis | 1 If testing for confounders was done or potential confounder variables were pruned |
|  | Presented SNPs | 2. SNPs significantly associated with the exposure were clearly listed, including their effect alleles, effect sizes and p-values.<br>1. SNPs associated with the exposure were listed but without complete information on effect alleles, effect sizes and p-values.<br>-1. SNPs were not listed |
| | P-value correction | 2: Applied<br>-1: When correction required was $< 10$ tests but not applied<br>-3: When correction required was $\geq 10$ tests but not applied<br>0: Not required |
|  | Was the study power considered? | 2: Yes<br>-1: No |
| Interpretation of results |  | 2: Results concluded appropriately according to statistical evidence<br>-2: Results not concluded appropriately according to statistical evidence |
| STROBE guidelines presented? |  | 1: Yes<br>0: No |

#### Study design

The study design category included an assessment of the study rationale (-1 to 2 points) by evaluating the quality of prior evidence supporting the association. Bidirectional studies, which test both the effect of the exposure on the outcome and the effect of the outcome on the exposure, were assigned 1 point because positive associations in both directions may indicate the presence of confounding factors (21). We also evaluated datasets based on several criteria: use of the most recent GWAS data (0 or 1 point), matching ancestries (-1 to 1 point), and absence of population overlap (-1 or 1 point). Matching ancestries was emphasized to minimize bias from ancestral differences in allele frequencies and linkage disequilibrium patterns (21). For studies comparing a multi-ancestry dataset to a single-ancestry dataset, 0 points were assigned if the specific ancestry of interest was not extracted or analyzed from the multi-ancestry dataset, as detailed in Table 1. In the same way, dataset independence was also assessed, as no participant overlaps between datasets reduces overestimation of genetic associations(22). Lastly, we evaluated whether replication was included in the study design, assigning 3 points for the inclusion of a replication strategy and -1 if replication was not conducted.

#### Statistical methods

The statistical methods evaluation included whether or not authors had addressed adherence to the three core MR assumptions (adequate strength of the instrumental variable, excluding genetic variants associated with known confounders of the relationship between exposure and outcome and considering whether the outcome is directly affected by the exposure) (6), appropriate p-value correction for multiple testing and whether or not power of the study was considered. One point was assigned if the selected SNPs demonstrated genome-wide significance (p < 5 × 10⁻⁸) or an F-statistic > 10. Additionally, we evaluated whether the SNPs were pruned for linkage disequilibrium (LD) using an R² threshold of < 0.1 (1 point), ensuring that only independent genetic variants were included in the instrumental variable. This step is important as it reduces bias in effect estimates (23). The second assumption states that no confounders should affect the causal relationship being assessed. This is challenging to test objectively, as all observational associations derived from epidemiological studies have unmeasured confounders(6, 24). Nevertheless, we assigned 1 point to studies that either adjusted for potential or known confounders or excluded SNPs associated with these confounders. The third assumption, requires that the genetic variant influences the outcome exclusively through the exposure (6). It is commonly tested using methods such as MR-Egger and MR-PRESSO, which detect outlier SNPs that may influence the outcome through pathways unrelated to the exposure. We did not directly score the use of them because most Mendelian randomization packages available in R incorporate these tests. However, an additional point was given to studies that conducted a mediator analysis alongside these MR methods (Table 1).

The statistical methods evaluation also comprised the presentation of the SNPs associated to the exposure along with effect sizes, effect alleles and p-values. Additionally, up to 3 points were given for appropriate multiple testing corrections (e.g., Bonferroni), and 2 points for considering study power in the methods or results sections.

#### Interpretation of results

We evaluated whether the results were interpreted correctly by considering several factors, such as the significance of the results after applying a Bonferroni correction when appropriate, if the authors accounted for evidence of high pleiotropy (e.g., a significant MR-Egger intercept or high distortion values in MR-PRESSO), and if the findings were consistent across multiple MR methods. The score for this criterion was of 2 or -2 points.

#### STROBE guidelines

1 point was assigned to studies that presented a table describing compliance with the MR-STROBE guideline criteria.

### Data Extraction and Statistical Analysis

Our data extraction included study year, definitions of exposure and outcome, dataset sources, MR methods, results, and score components (Supplementary Table 1). We used Shapiro-Wilk tests for score distribution and examined trends in article quality using linear regression, with significance set at p < 0.05.

## Results

PMID and summary information of the 86 articles, published between 2016 and March 2024, is presented in the Supplementary Table 1. Among them, 70 focused on urate as either the exposure or outcome variable (Figure 2). In 59 of the 70 studies, urate was the exposure, with 27% (16/59) reporting a causal relationship. Common phenotypes found to be causally associated with increased serum urate included coronary heart disease, hypertension, heart failure, and myocardial infarction (Table 2). Of the 59 studies, 34 conducted bidirectional analyses, while 25 focused solely on urate as the exposure and 11 as the outcome (Figure 3). Conversely, 31 studies used urate as the outcome variable of which 22 (71%) found causal associations with BMI, fasting insulin, HDL cholesterol, and triglycerides (Table 2).

**Figure 2.**
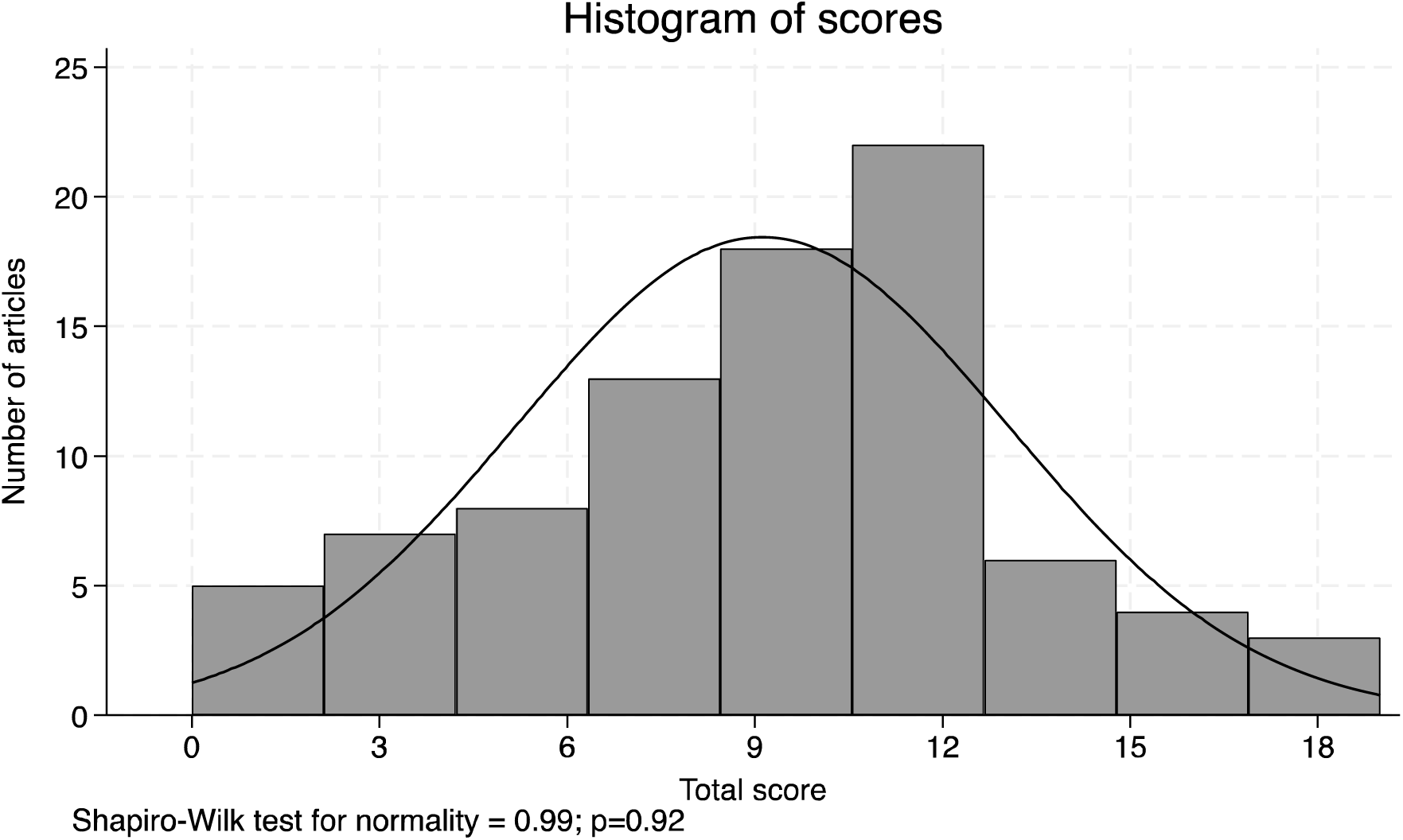
Histogram of scores.

**Figure 3.**
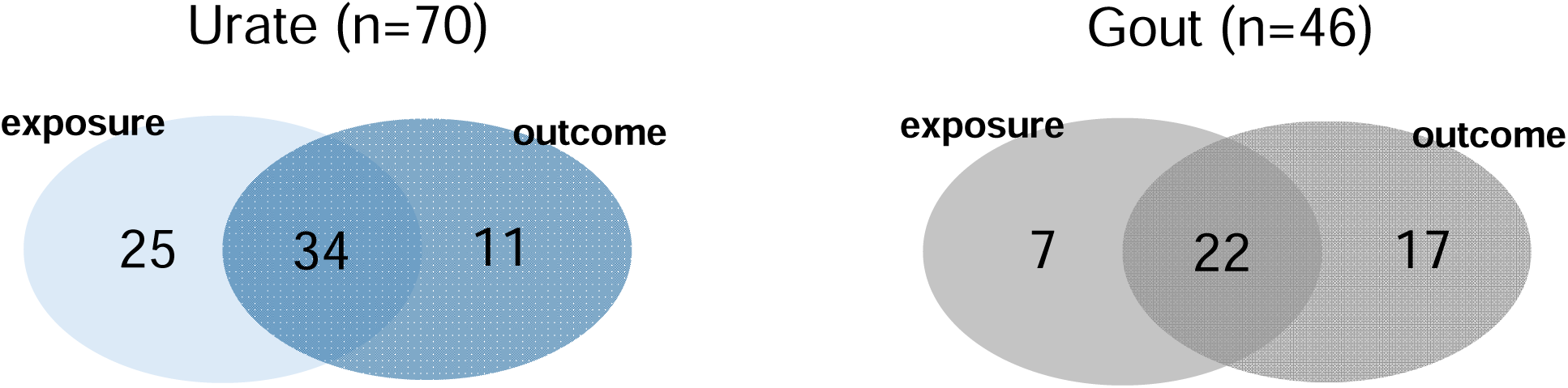
Number of studies using urate and gout as variables.

**Table 2.** Most frequently studied phenotypes in gout and urate MR studies.

| Exposure | Outcome | N<br>articles | Articles that found association |  | Articles that found no association |  |
| --- | --- | --- | --- | --- | --- | --- |
|  |  |  | Mean<br>score | Articles | Mean<br>score | Articles |
| Urate → trait |  |  |  |  |  |  |
| Urate | Coronary heart<br>disease | 8 | 9.3 | 75, 176, 200 | 2.3 | 12, 100, 129, 131, 136 |
| Urate | Hypertension | 6 | 2.1 | 46, 78 | 10.5 | 65, 97, 129, 131 |
| Urate | BMI | 4 | - | - | 1.9 | 5,31,60,65 |
| Urate | Heart failure | 4 | 9.1 | 62,78 | 2.1 | 1,26 |
| Urate | CKD | 3 | - | - | 10.7 | 97,41,137 |
| Urate | Gut microbiota | 4 | - | - | 6.25 | 2,30, 43, 199 |
| Urate | Myocardial<br>infarction | 3 | 14 | 75 | 10 | 129, 131 |
| Urate | Fasting insulin | 3 | - | - | 12.3 | 65, 91,99 |
| Gout → trait |  |  |  |  |  |  |
| Gout | Coronary heart<br>disease | 2 | 5 | 78, 200 | - | - |
| Trait → Urate |  |  |  |  |  |  |
| BMI | Urate | 7 | 9 | 5,15,31,60,65,93, 139 | - | - |
| Coffee | Urate | 4 | 7 | 9, 38 | 8 | 73,106 |
| Gut<br>Microbiota | Urate | 4 | - | - | 6.25 | 2,30, 43, 199 |
| Fasting Insulin | Urate | 3 | 12.3 | 65, 91,99 | - | - |
| Waist/Hip<br>ratio | Urate | 3 | 9 | 31 | 9 | 139, 65 |
| HDLc | Urate | 3 | 11.3 | 65,93,102 | - | - |
| TG | Urate | 3 | 11.3 | 65,93,102 | - | - |
| T2DM | Urate | 2 | 9 | 99 | 11 | 65 |
| Trait → Gout |  |  |  |  |  |  |
| Tea intake | Gout | 4 | 10 | 26,215 | 3 | 16,211 |
| BMI | Gout | 3 | 9.67 | 31,65,93 | - | - |
| Coffee | Gout | 2 | 5.5 | 73,142 | - | - |
| Blood<br>pressure | Gout | 2 | 13 | 65,198 | - | - |
| Gut<br>microbiota | Gout | 2 | - | - | 9.5 | 30,43 |

For gout-related MR analyses, 46 studies were included. Of these, 29 used gout as the exposure variable (Figure 3), with only one reporting a causal relationship, which was with coronary heart disease (Table 2). Additionally, 39 studies investigated potential causes of gout (Figure 3) of which 14 reported causal associations, most commonly with tea intake, coffee, BMI, and high blood pressure.

The scores assigned to the 86 studies ranged from 0 to 19, with a mean of 9.1 and median of 11, and were normally distributed (Shapiro-Wilk: 0.99, p=0.92) (Figure 2). The following paragraphs describe the results of our scoring criteria in detail.

The first aspect evaluated in the study design was the rationale. We assessed the plausibility of the studied phenotypes by reviewing prior evidence of the associations. Strong observational evidence, such as findings from large observational studies or small clinical trials supporting the hypothesized association, earned 2 points. Mixed evidence or findings from small observational studies earned 1 point, while evidence based on fewer than five small studies received -1 point (Table 1). Overall, 55 phenotypes (64%) had a strong rationale, 29 (34%) showed mixed or weak evidence, and 2 (2%) were given the lowest score.

Another aspect evaluated in the study design was dataset quality. The most commonly used urate dataset was the GWAS study published of 110,347 individuals by Köttgen et al.(25) in 2013 (n=44), followed by the GWAS dataset of 457,690 individuals published by Tin et al.(26) in 2019 (n=20)(Table 3). Overall, 34% of the studies used outdated datasets, meaning that they used the Köttgen et al. dataset instead of the four times larger Tin et al. dataset published in 2019 when the Tin et al. dataset was available. The main ancestry in the two datasets was European, which was also the most studied ancestry among the MR studies. Table 3 summarizes the datasets and ancestries used in the articles included in our review.

**Table 3.** Datasets used by MR studies.

| Dataset | Ancestry | Year | Urate sample size | Gout sample size (cases / controls) | Freq (%) | PMID |
| --- | --- | --- | --- | --- | --- | --- |
| Köttgen | European | 2013 | 110347 | 2115 / 67259 | 44 (51.16%) | 23263486 |
| Tin | European | 2019 | 288649 | 13179 / 750634 | 20 (23.26%) | 31578528 |
| Japan Biobank | East Asian | 2019 | 109029 | 3053 / 4554 | 6 (6.98%) | 32238385 |
| UK Biobank | European |  | NA | 6542 / 456391 | 12 (13.95%) |  |
| Sakaue | European + East Asian | 2021 | 343836 | - | 2 (2.33%) | 34594039 |
| FinnGen | European |  | - | 3576 / 147221 | 8 (9.3%) |  |
| UK Biobank | African | 2021 | 6206 | - | 1 (1.16%) |  |
| Taiwan Biobank | East Asian | 2008 | 34831 | - | 1 (1.16%) | 18370851 |
| Nakatochi | East Asian | 2019 | 121745 | - | 1 (1.16%) | 30993211 |
| Kolz | European | 2009 | 28141 | - | 1 (1.16%) | 19503597 |
| Huffman | European | 2015 | 42569 |  | 2 (2.33%) | 25811787 |
| White | European | 2016 | 166486 | - | 2 (2.33%) | 26781229 |
| Leon-Mimila | Hispanic | 2013 | 1073 adults, 1080 children | - | 1 (1.16%) | 23950976 |
| Dönertaş | European | 2021 | - | 488295 | 1 (1.16%) | 33959723 |
| Zhou | European + East Asian | 2022 | - | 30549/ 1039290 | 1 (1.16%) | 36777996 |

We also considered if the exposure and outcome datasets studied the same ancestry. 8% of the studies did not use datasets with matching ancestries and 5% used a dataset with a mixed-ancestry dataset for comparison against another with a single ancestry. Additionally, we reviewed whether the datasets being compared had participants that overlapped between the exposure and outcome datasets, and we found that only 51% of the studies used independent datasets.

The statistical methods category contained the evaluation of MR assumptions. Of the studies reviewed, 9 (17%) satisfied all our criteria addressing the three assumptions and received the highest scores in those criteria. Additionally, 66% of the studies satisfied the first assumption, also known as relevance; while 50% tested for confounders.

Regarding the power of the study, we assigned 2 points to studies that addressed it in the methods or the results section or if they actually calculated it anywhere in the manuscript. 59% fulfilled this criterion.

Another criterion in our scoring system focused on the interpretation of results. Our evaluation considered whether the conclusions presented in the studies were consistent after the p-value was corrected for multiple testing. Of the 86 studies, 56 (65%) reported a causal association. However, 12 studies (14%) drew incorrect conclusions about the causal association due to the lack of necessary p-value Bonferroni correction. Also, we assessed whether the results were replicated in independent datasets and found that only 12 studies (14%) conducted a replication analysis.

### Score trends per year and place of origin

An analysis of mean article scores by year of publication revealed a significant downward trend, indicating a decrease in average article quality over time (β= -0.29, p= 0.0009). Additionally, we compared the scores before and after the publication of STROBE guidelines for MR (20)and found no difference in the scores (p=0.58). When analyzing the score variability per year, it is of note that article quality is becoming more diverse. The highest scores showed a positive trend (β = 0.74, p < 0.00001), indicating an increase over time, while the lowest scores declined at a faster rate (β = -1.25, p < 0.00001), suggesting a widening gap in the quality of MR studies in urate and gout (Figure 4).

**Figure 4.**
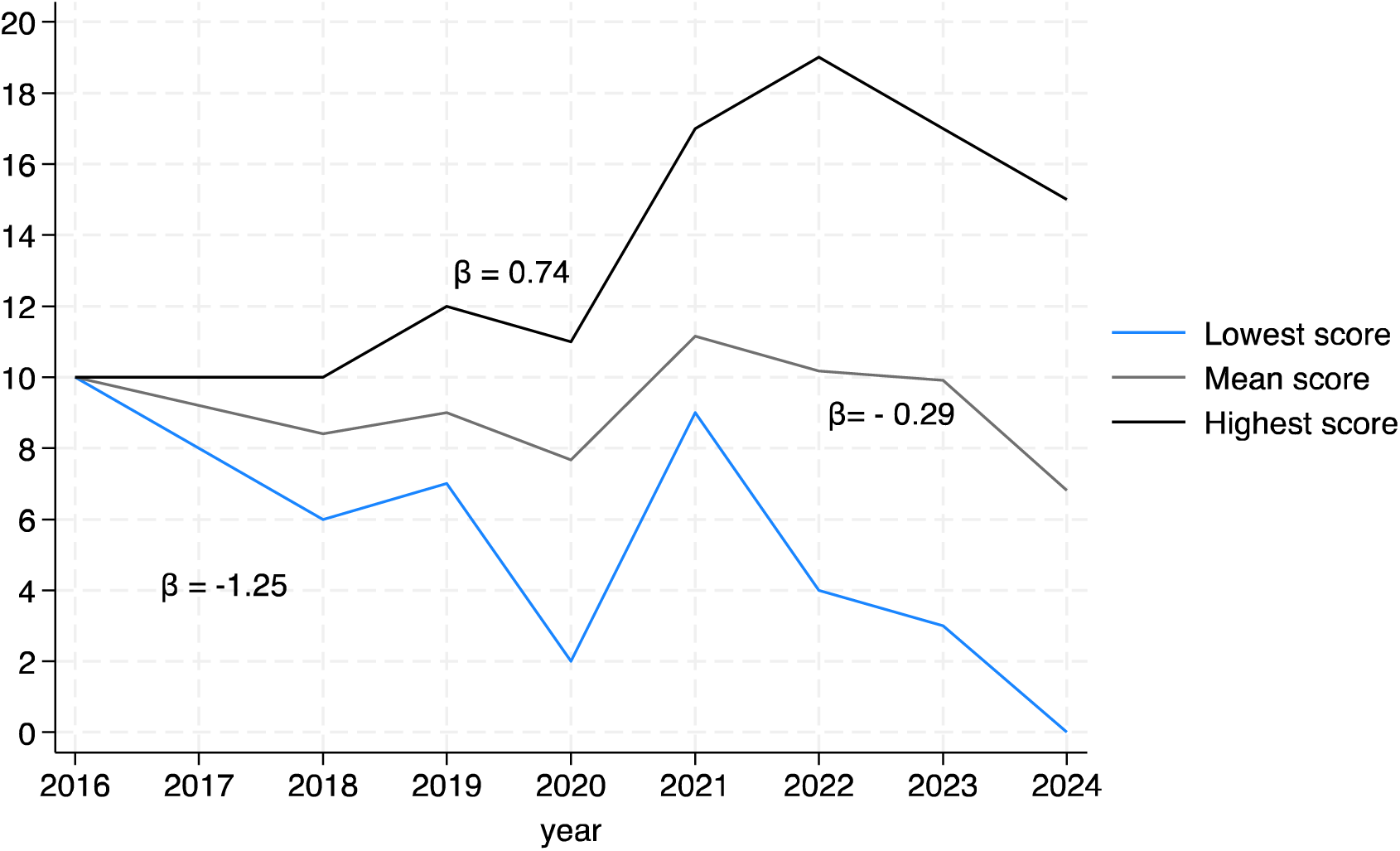
Trends in scores by year.

Articles were also categorized by the first author’s country of origin. Most were from China (70%), followed by the USA (6%) and UK (5%). For comparative purposes, articles were grouped by continent. Asia had the lowest mean score (8.9 ± 0.5), while Oceania had the highest (10 ± 0.6). Notably, Asia exhibited the widest range of scores, reflecting both the lowest and highest scoring articles (IQR = 5), while Oceania had the least variability (IQR = 2) (Table 4). No significant differences in scores were observed between continents (p=0.8). Over time, the geographic distribution of studies shifted, with considerably more MR studies recently originating from Asia and fewer from other continents (Figure 5) - since the beginning of 2022 96% of MR studies were from Asia.

**Figure 5.**
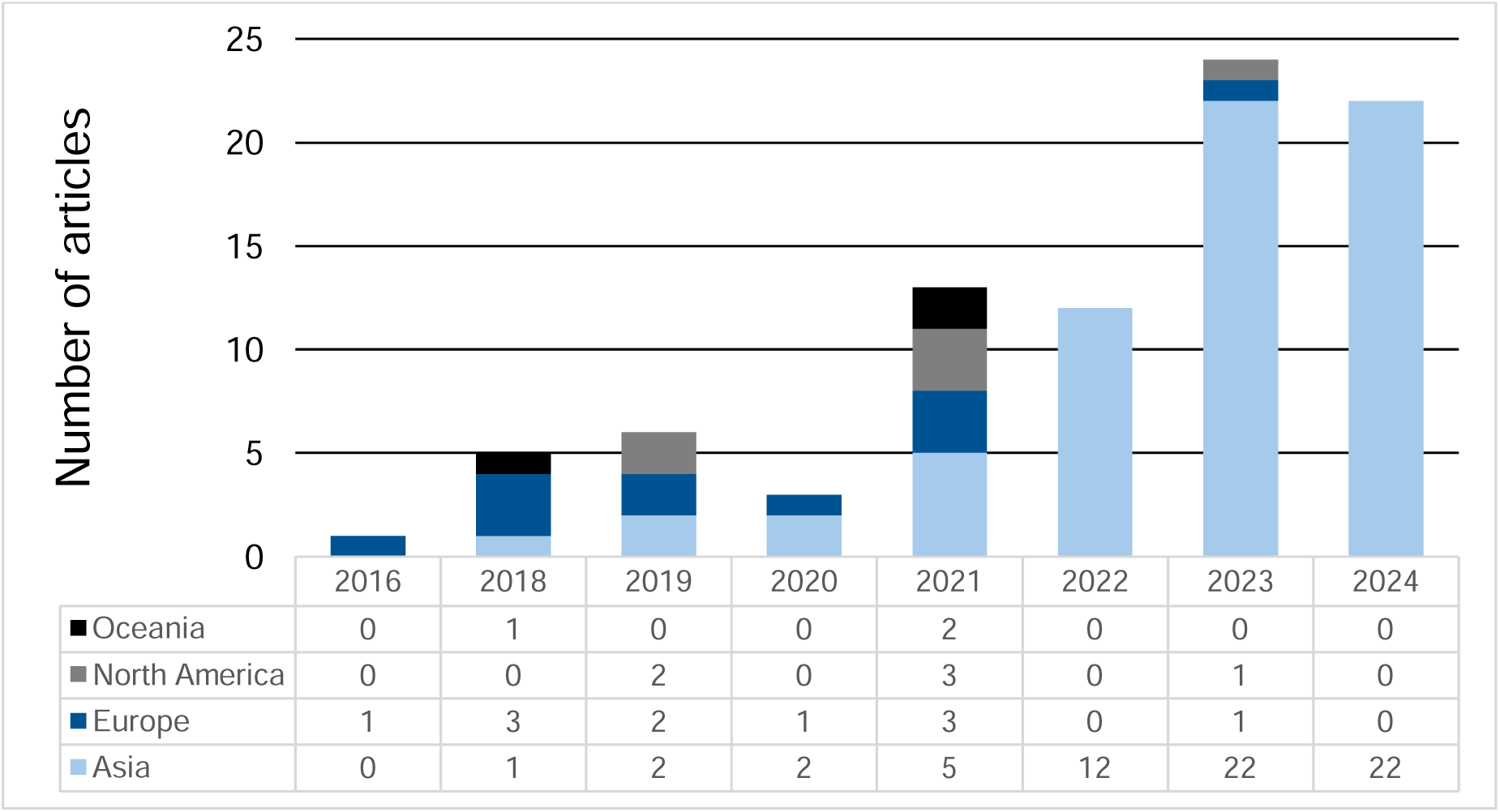
Continent of origin over time.

**Table 4.** Summary of mean scores and variability by continent.

| Continent | Mean | IQR |
| --- | --- | --- |
| Asia | 8.9 ± 0.5 | 5 |
| Europe | 9.8 ± 0.7 | 4 |
| North America | 9.5 ± 2 | 3 |
| Oceania | 10 ± 0.6 | 2 |
| Total | 9.1 ± 4 | 4 |

IQR= Interquartile Range

## Discussion

Over the past 12 years, we found a steady increase in 2-sample Mendelian randomization (MR) studies, rising from one in 2016 to 22 in 2022. However, the quality of these studies varies widely, with scores ranging from -2 to 19 and an average score of 9.1 (Table 4). This growth is largely driven by the expanding availability of GWAS and the relative ease of conducting MR analyses (24, 27). While this rise has certainly contributed to the understanding of causal relationships, it has also led to studies with poor rationale, skewed estimates and weak instrumental variables, results lacking robustness, and inaccurate conclusions (6, 24).

To address inconsistencies in study quality, guidelines such as the 2021 STROBE-MR guideline have been introduced(20, 24, 28). These guidelines focus on increasing the quality reporting of MR studies, rather than a framework to increase quality of an MR study per se. However, the overall quality of published articles has not improved, likely due to a lack of adherence as only 11% of articles published after 2021 reported following the guideline. Studies that followed it demonstrated higher quality, with compliant articles averaging a score of 10.1, compared to the overall post-2021 average of 8.8.

The lowest-scoring articles in our analysis, each receiving a score of 0, revealed significant gaps in study design. Both studies lacked result replication, confounder testing, adequate p-value corrections, and an assessment of statistical power. None of the studies presented the specific SNPs used or their association values with the exposure. The first study, focused on sepsis and gout/urate (article 6 in Supplementary Table 1), used a dataset with European and Japanese ancestry for the exposure and a European ancestry dataset for the outcome. The second study, on sex hormones, breast cancer, and gout (article 202 in Supplementary Table 1), used datasets with European ancestry for most phenotypes but included an East Asian ancestry dataset for the urate trait, resulting in mismatched datasets.

In contrast, the highest-scoring articles received scores of 19 and 17. Article 62 (Supplementary Table 1), which reported a causal relationship between urate and heart failure, achieved the highest score overall. This study satisfied all the core assumptions of MR and got the highest score in most criteria except for presentation of the STROBE guidelines. Articles 21 and 91 (Supplementary Table 1) tied for the second-highest score. Article 21 found that metformin had a preventive effect on high urate levels but not on gout, while article 91 identified hyperinsulinemia as a cause of elevated urate levels. One of the weaknesses we identified on article 21 was that it was unidirectional. Also, they failed to use the latest dataset for gout and urate and did not present adherence to the STROBE guideline. Similarly, article 91 fulfilled most criteria but lacked a mediator analysis and also did not include the STROBE guidelines. This last study was published only 1 month after the publication of the guideline, which may explain the omission.

Ensuring that MR assumptions are met is essential for valid results. In our review, the relevance assumption was met by 96% of studies. However, only 50% addressed the independence assumption by explicitly discussing confounders. The third assumption requires that the genetic instrumental variables influence the outcome solely through the exposure. The exclusion restriction assumption may not be met in cases where exposures are also affected by environmental factors (e.g., education, physical activity, vitamin D levels). Eight studies (9%) violated this assumption due to implausible exposures (see Supplementary Table).

Additionally, a common challenge in two-sample MR studies is overlapping datasets, which cause inflation of effect estimates (29, 30). This issue is particularly relevant for non-European ancestry studies, where limited large datasets often necessitate overlap, whereas European ancestry studies benefit from broader dataset availability. Most studies in our analysis relied on datasets from large GWAS consortia, which enhances study power but can lead to participants overlapping between exposure and outcome datasets. Among the 86 studies reviewed, only 52% of the studies used fully independent datasets.

Our analysis also demonstrates how the 2021 STROBE-MR guideline(20) and our scoring system complement one another while serving distinct purposes. The STROBE-MR guideline is a comprehensive checklist to guide authors during study preparation, which attempts to standardize all aspects of the manuscript including statement of objectives, participant eligibility criteria and the software used for the analysis. In contrast, our scoring system aims to quantitatively evaluate the quality of MR studies after their completion, focusing on specific key aspects like study design, SNP selection and result interpretation (Table 1). For example, while STROBE included descriptions of MR assumptions and contemplated many types of sensitivity analyses, our scoring system approaches the assumptions through specific criteria, such as p-value thresholds for SNP selection and explicit confounder testing. Similarly, while STROBE include a category for sensitivity analyses that include comparisons of effect estimates from different methods, independent replication, bias analyses, validation of instruments, or simulations(20), we focused only on replication for simplicity and because replication is the superior metric. Additionally, both systems assess generalizability differently. Our scoring system focuses on matching dataset ancestries and ensuring dataset independence, while STROBE approaches generalizability in a less specific manner and includes discussions on biological mechanisms. These two frameworks serve distinct purposes at different stages of MR studies. The STROBE-MR guideline is intended for researchers preparing manuscripts, whereas our scoring system is a practical tool for reviewers or readers who need to assess the quality and validity of completed MR studies.

Others have also identified the impact of the current load of two-sample MR studies on the medical literature. Stender et al.(31), in a paper entitled “Reclaiming mendelian randomization from the deluge of papers and misleading findings”, pointed out that public availability of GWAS summary statistics has prompted “an explosion of low-quality two-sample mendelian randomization studies”. They state that “These studies add minimal – if any – value and overwhelm reviewers and journals.”. Stender at al. also advise editors to reject without review two-sample MR papers that only report the MR findings per se with no additional supporting evidence. We fully support these views.

Table 2 summarizes the most common phenotypes linked to urate and gout identified in our review. BMI was consistently identified as a causal factor for elevated serum urate in populations of European ancestry, with supporting studies averaging a score of 9. These findings align with those from older MR studies using different MR techniques that support a causal relationship between BMI and urate, such as the ones performed by Lyngdoh, et al (32), Palmer, et al (33)and Oikonen, et al (34). Further evidence comes from a randomized controlled trial involving 235 patients, which found that weight loss led to decreased urate levels, regardless of diet type (35). This example demonstrates that MR results in urate and gout can be applicable and informative in clinical settings. However, it is important to note that the effect of BMI on urate levels is relatively small and, clinically, the use of urate-lowering therapy to manage gout is considerably more effective.

Other phenotypes with more than one study supporting a causal association included gout as a potential cause of coronary heart disease (CHD). However, the number of two-sample Mendelian randomization (MR) studies was limited (n=2), and their mean quality score was below the overall average (Table 2). This association may be mediated by hyperuricemia, as urate is known to act as a pro-oxidant under hyperuricemic conditions and in the presence of chronic diseases such as metabolic syndrome, chronic heart failure, and chronic kidney disease. In these settings, urate contributes to endothelial dysfunction by reducing nitric oxide production and activating the renin-angiotensin system, processes that promote atherosclerosis (36). However, in the clinical setting, the role of urate on cardiovascular disease is not yet fully understood. Results from the CARE trial (2018), that compared febuxostat and allopurinol, showed that, even though febuxostat had a larger percentage of patients with urate levels under 5.0mg/dL, there was no difference in cardiovascular mortality (37). Furthermore, a recent one-sample MR study found no significant effect of lowering urate levels through xanthine dehydrogenase-related SNPs on the risk of ischemic heart disease (38). Similar to the current literature, our review yielded mixed results regarding the role of urate in CHD. Three studies with a mean score of 8 supported this association, while five studies with a mean score of 2.3 did not (Table 2).

Three studies investigated a causal relationship between blood lipids and serum urate levels, consistently identifying high-density lipoprotein cholesterol as a preventive factor against elevated serum urate and triglycerides as a causal factor for increased serum urate (Table 2). However, older studies, including one-sample MR analyses, have shown mixed findings, with some studies favoring the direction of blood lipids influencing serum urate, while others support the reverse relationship (39, 40, 41). This variation across studies raises concerns about horizontal pleiotropy and suggests that the observed associations may be influenced by confounding factors or shared pathways.

Similarly, the effect of urate-lowering drugs on blood pressure has also yielded mixed results(42, 43, 44). MR studies, along with our analysis of two studies (Table 2), identified blood pressure as a potential cause of gout. It would be valuable to investigate whether managing blood pressure in patients with risk factors for gout or administering antihypertensive medications to those with hyperuricemia could reduce the risk of developing gout.

While MR has advanced our understanding of causal relationships, many studies fall short in meeting core assumptions, using independent datasets, or conducting replication analyses. BMI stood out as a consistent causal factor for hyperuricemia, supported by both MR and clinical evidence. Our scoring system provides a practical tool for evaluating MR study quality, helping researchers and clinicians identify strengths and weaknesses in design, methodology, and interpretation.

## Data Availability

All data produced in the present work are contained in the manuscript

---

Supplementary table 1. (Excel file)

